# Poor glycemic control and associated factors among diabetic patients in Ethiopia; A Systemic review and meta-analysis

**DOI:** 10.1101/19004986

**Authors:** Berhane Fseha Teklehaimanot, Abadi kidanemariam Berhe, Gebrehiwot Gebremariam Welearegawi

**Affiliations:** Lecturer in Epidemiology & Biostatistics,Department of public health, College of Medicine and Health Sciences, Adigrat University, Tigray, Ethiopia; Assistant professor in Department of Nursing, College of Medicine and Health Sciences, Adigrat University, Tigray, Ethiopia; Lecturer in reproductive health, Department of Public Health, College of Medicine and Health Sciences, Adigrat University, Tigray, Ethiopia

**Keywords:** Poor glycemic control, diabetes, Fasting blood glucose, glycated hemoglobin, Ethiopia

## Abstract

**Introduction:** The major global public health problems now days are diabetes especially the burden is high in low income countries including Ethiopia due to the limited resource for screening and early diagnosis of the diabetes. To prevent diabetic complications including organ damage and micro vascular complications blood glucose level should be maintained at an optimum level. However there was no pooled national picture on poor glycemic control and its associated factors.

**Methods:** Different data base searching engine including PubMed, Google scholar, the Cochrane library, MEDLINE,, HINARY and African journal online (AJOL) were used. The Joanna Briggs Critical Appraisal Tools and Newcastle Ottawa scale for assessing the quality of cross sectional studies were used for quality assessment. The meta-analysis was conducted using STATA 14 software. *I*^*2*^ statistic and egger weighted regression were used to assess heterogeneity and publication bias.

**Results:** A total of 134 studies were identified from different database searching engines and other sources. After removing for duplication, absence of abstract and review of the full text 12 studies were including in the meta-analysis. The pooled prevalence of poor glycemic control among diabetic patients in Ethiopia is 64.72% with 95% confidence interval 63.16-66.28%. The sub group analysis of poor glycemic control among diabetic patients in different region of the country shows consistent and high prevalence of poor glycemic control ranging from 62.5% in Tigray region to 65.6% in Oromia region of the country. Residence, dyslipidemia and diet adherence were significantly association with poor glycemic control among diabetic patients in Ethiopia.

**Conclusion:** The prevalence of poor glycemic control among diabetic patients was high in Ethiopia and consistent across different regions of the country. The most important factors associated with poor glycemic factor among diabetic patients were being rural residence, having dyslipidemia and not adhering to dietary plan.

## Introduction

Diabetes is chronic disease occurred due to increased blood glucose level because of the body cannot produce at all or secrets in sufficient insulin hormone or not use it effectively. Hence, the nonexistence of insulin or the cell is not sensitive to use insulin leads to increased blood glucose level which is the hallmark of diabetes(1). Globally 425 million adult population have diabetes and an estimated of 15.5 Millions of them are living in Africa and it is projected that by 2040 about 34.2 million are expected to be diabetic(2). One of the major global public health problems now days are diabetes especially the burden is high in low income countries including Ethiopia due to the limited resource for screening and early diagnosis of the diabetes (3).

To prevent diabetic complications including organ damage and micro vascular complications the blood glucose level should be maintained at an optimum level because poor glycemic control leads to diabetic keto acidosis, cognitive impairment, immune dysfunction and hospital admissions due to May complications. The magnitude of poor glycemic control is high throughout the world and specifically in Ethiopia greater than 50% of the diabetic patients does not had optimum blood glucose level(4–7).

Several primary Studies have been conducted in Ethiopia on the magnitude of poor glycemic control and their associated factors among diabetic patients. However the result of these studies were not consistent on the prevalence of poor glycemic control and even the factors that have significant association with poor glycemic control among diabetic patients varies across the studies. Moreover, there is no a study that shows the national pooled prevalence of poor glycemic control and contributing factors as well.. Therefore, this systemic review and meta-analysis on poor glycemic control and associated factors in Ethiopia are filling these gap and will generating new attention on the significant contributing factors to maintain the optimum blood glucose level among diabetic patients in Ethiopia.

## Methods

### Study design and search strategy

A systematic review and meta-analysis on the poor glycemic control and associated factors among diabetic patients in Ethiopia has been conducted. Different data base searching engine including PubMed, Google scholar, the Cochrane library, MEDLINE, HINARY and African journal online (AJOL) were used. Besides, grey literature and the reference list of former studies were also searched for extra articles. The Boolean operators was used to search the articles by using the following key and medical subject hedging terms separately and in combination “glycemic control” “glycemic control”, “blood glucose control”, “fasting blood glucose”, “glycated hemoglobin”, prevalence, magnitude, “associated factor”, predictors, Ethiopia. Language was limited to be only English. From May 2019 up to July 2019 was the period used to search the studies.

### Study selection and eligibility criteria

#### Study selection

Selection of articles for the systemic review and meta-analysis were if the study conducted in Ethiopia and studies reporting poor glycemic control and/or the associated factors. Two independent researchers assessed the abstract and full text of all articles after preliminary screening of the studies for their title. A third researcher has been resolved the disagreement between the two researchers. A total of 134 studies were found from database searching and grey literatures (figure 1).

**Figure 1.**
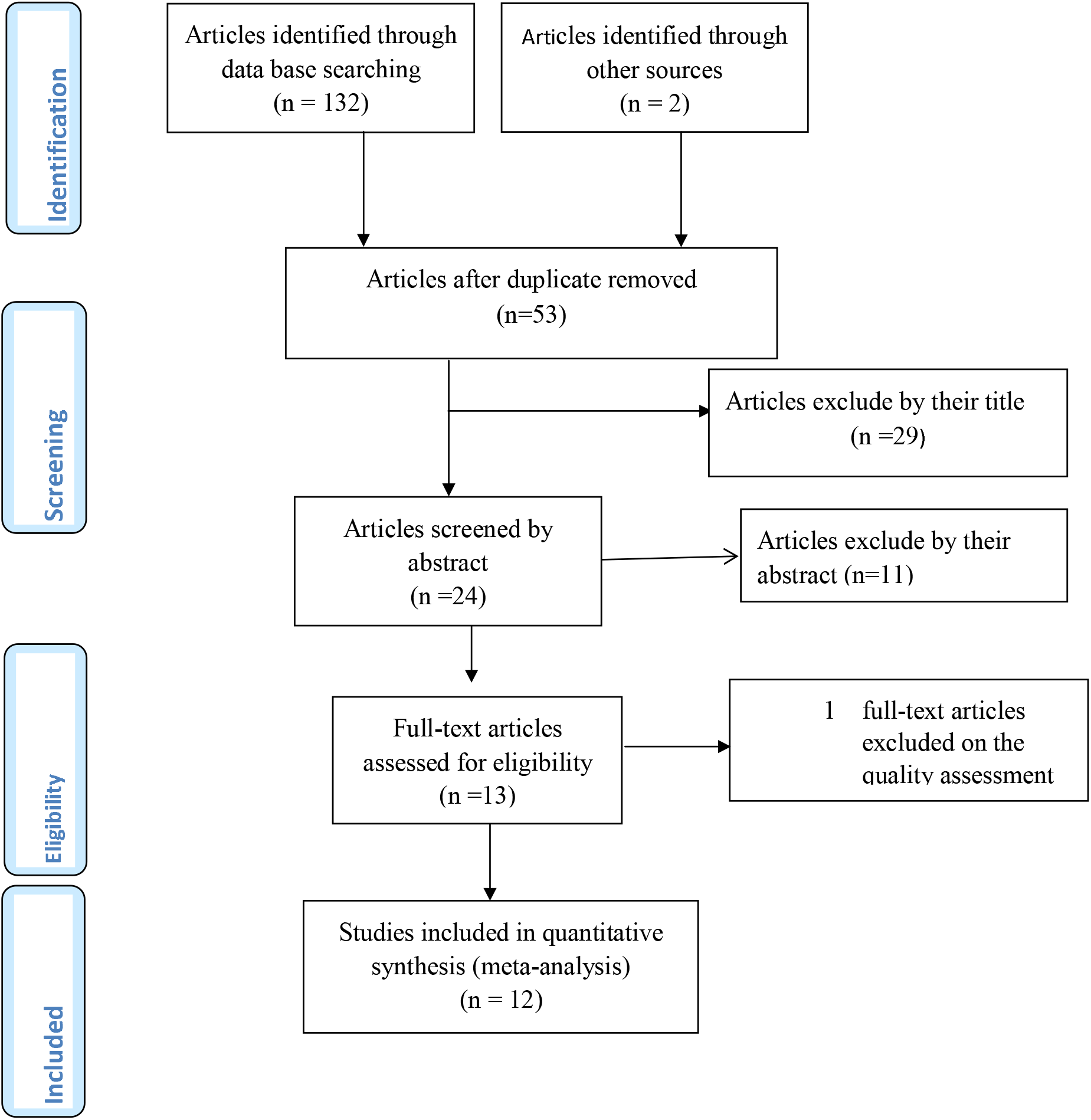
Flow chart diagram showing the selection of articles for systemic review and meta-analysis of poor glycemic control and associated factors among diabetic patients in Ethiopia 2019.

### Eligibility criteria

#### Inclusion

✓ All articles published until the end of July 2019
✓ Only researches measured the poor glycemic level and/or associated factors among diabetic patients published and grey literatures were included
✓ Fasting blood glucose, oral glucose tolerance test and hemoglobin a one c(Ha1c) measurement for poor glycemic control according to the standard cut off point were included
✓ For duplicated articles the most recent were used
✓ Only English language published articles were used

#### Exclusion

✓ Studies used random blood glucose level to determine poor glycemic control were excluded
✓ Poor glycemic level that do not adhere to the standard world health organization definition and international diabetic federation were excluded.
✓ Article that are rated as low quality assessment was rejected.
✓ Studies that have response rate of <80% were excluded

#### Data extraction

Data extraction form has been developed by two researcher in excel spread sheet format and appraised by the third author. The data abstraction includes author, year of study, year of publication, study area(region of the country), study design, study setting(hospital/community based), sampling technique, sample size, response rate, outcome measured by(fasting blood glucose/ HbA1c), prevalence of poor glycemic control, age, gender, residence(urban/rural), monthly income, educational status, occupation, regular follow up, serum lipid level, presence of comorbidity, duration of disease, physical exercise, medication adherence, diet adherence, type of diabetes(type 1/type 2), medication(insulin, oral and combined), and body mass index (BMI) of the study participant were extracted. In addition to the total prevalence of poor glycemic control and sub categories by region of the country and outcome measured by fasting blood glucose or hemoglobin HbA1c has been extracted.

#### Operational definition of outcome measures

Poor glycemic control is the level of blood glucose rages out of the normal value that is Fasting blood glucose measurement greater than 130mg/dl and less than 70 mg/dl and glycated hemoglobin(Ha1c≥ 7%).

#### Study quality and critical appraisal

Quality of the studies was assessed by the Joanna Briggs Critical Appraisal Tools and Newcastle Ottawa scale for assessing the quality of cross sectional studies in meta-analysis(8). Parameter for quality assessment contains sampling procedure, representativeness in terms of their sample size, response rate, and blood glucose measured by standard measurement tool.

#### Heterogeneity and publication bias

Cochranes Q test which shows the presence of heterogeneity among studies and I^2^ statistic were used to test heterogeneity. The estimate of variation in effect estimate that is due to heterogeneity rather than sampling error or chance differences was evaluated by I^2^ statistic. Hence, the presence of heterogeneity was checked by Cochranes Q test (p<0.10 indicates statistically significant heterogeneity as cut off point) and the value of I^2^ test 25%, 50%, 75% indicates low, medium and high heterogeneity among studies.

To assess for publication bias egger weighted regression and Begg rank correlation test (p< 0.05 were considered as statistical significant publication bias) method used.

#### Statistical analysis

The data entry was conducted on Microsoft excel spread sheet for windows 2010 and exported to STATA version 14 statistical software for analysis. Descriptive summary of the original studies were conducted and presented in forest plot, tables and texts. Fixed effect model was used to compute the pooled prevalence with 95% confidence interval of poor glycemic control since there was no heterogeneity among the articles and random an fixed effect model was computed to identify the factors that had significant association with poor glycemic control among diabetic patients in Ethiopia.

#### Subgroup analysis

Subgroup analysis were conducted by region of the country, outcome measured by fasting blood glucose/ or hemoglobin a1c and pooled prevalence of the factors was displayed by forest plot with the effct size and 955 confidence intervals.

## Result

### Identified studies

A total of 134 studies were identified from different database searching engines PubMed, Google scholar, the Cochrane library, MEDLINE, HINARY, and African journal online (AJOL). After removing for duplication, absence of abstract and review of the full text does not have the poor glycemic control measurement only 13 studies were eligible to include for meta-analysis. We exclude one article based on the quality assessment result. Finally 12 articles have been included in the meta-analysis. Figure 1

### Study characteristics

In the meta-analysis total of 12 studies were included. Majority 5(41.67%) of the study were from Amhara region of the country, three (25%) from Oromia, two (16.67%) from Tigray and two (16.67%) from Addis Ababa the capital city of Ethiopia. All of the studies were cross sectional in study design and the largest sample size was in Addis Ababa which was 423 and the smallest sample size was in Amhara region (Table 1).

**Table 1:** Distribution of selected studies for the systemic review and meta-analysis of poor glycemic control and associated factors among diabetic patients in Ethiopia, 2019

| S. № | Author(publication year) | Region | Study design | Study setting | Sample size | Resposerate(%) | Blood glucose measured by | Poor glycemic control(%) |
| --- | --- | --- | --- | --- | --- | --- | --- | --- |
| 1 | Abebe, S. et al (2015) | Amhara | cross sectional | hospital | 407 | 100 | HbA1c | 64.7 |
| 2 | Ayele, A. &Tegegn, H.(2019) | Amhara | cross sectional | hospital | 278 | 98.9 | FBG | 57.1 |
| 3 | Cheneke, W. et. Al (2016) | Oromia | cross sectional | hospital | 148 | 100 | HA1C | 59.5 |
| 4 | Demoz, G. et.al (2019) | Addis ababa | cross sectional | hospital | 423 | 84.4 | FBG | 68.3 |
| 5 | Fasil et al (2018) | Amhara | cross sectional | hospital | 367 | 100 | FBG | 60.5 |
| 6 | Fiseha, T et.al (2018) | Amhara | cross sectional | hopspital | 384 | 100 | FBG | 70.8 |
| 7 | Fseha, B. (2017) | Tigray | cross sectional | hospital | 200 | 100 | FBG | 63.5 |
| 8 | Gebre-Yohannes, A. &Rahlenbeck, S. I. (1997) | Amhara | cross sectional | hospital | 102 | 100 | HA1C | 77 |
| 9 | Kassahun, T. et.al (2016) | Oromia | cross sectional | hospital | 325 | 95 | FBG | 70.9 |
| 10 | Mideksa et.al (2018) | Tigray | comparative cross sectional | hospital | 336 | 100 | HA1C | 61.9 |
| 11 | Shimels, T. et.al (2018) | Addis ababa | cross sectional | hospital | 414 | 87.2 | FBG | 60.3 |
| 12 | Yigazu, D. &Desse, T. (2017) | Oromia | cross sectional | hospital | 74 | 100 | FBG | 59.2 |

**Table 2:** Distribution of factors associated with poor glycemic control in the selected studies in Ethiopia, 2019

| S. No | Author(publication year) | Number of Poor glycemic control |  |  |  |  |  |  |  |  |  |  |  |  |  |  |
| --- | --- | --- | --- | --- | --- | --- | --- | --- | --- | --- | --- | --- | --- | --- | --- | --- |
|  |  | age |  |  | Sex |  | Residence |  | Income |  | Occupation |  | Educational status |  | comorbidity |  |
|  |  | Age >65 | Age 25-65 | Age <25 | male | female | Urban | Rural | poor | rich | empl oyed | unemp loyed | Formal education | No formal education | Yes | no |
| 1 | Abebe, S. et al (2015) | 32 | 195 | 25 | 125 | 128 | 215 | 38 | 167 | 52 |  |  |  |  | 98 | 155 |
| 2 | Ayele, A. &Tegegn, H.(2019) | 24 | 133 |  | 83 | 74 | 99 | 58 | 90 | 38 |  | 33 | 47 | 100 |  |  |
|  | Cheneke, W. et. Al (2016) |  |  |  | 50 | 38 |  |  |  |  |  |  | 57 | 31 |  |  |
| 4 | Demoz, G. et.al (2019) | 96 | 148 |  | 74 | 170 | 206 | 38 |  |  | 135 | 109 | 171 | 33 | 195 | 49 |
| 5 | Fasil et al (2018) | 30 | 162 | 30 | 106 | 116 | 157 | 65 |  |  | 139 | 83 | 138 | 99 |  |  |
| 6 | Fiseha, T et.al (2018) |  |  |  |  |  | 181 | 91 |  |  | 145 | 126 | 150 | 121 |  |  |
| 7 | Fseha, B. (2017) | 5 | 119 | 2 | 83 | 43 | 88 | 38 | 65 | 24 |  |  | 82 | 45 | 44 | 82 |
| 8 | Kassahun, T. et.al (2016) |  |  |  |  |  |  |  |  |  | 172 | 47 | 138 | 81 |  |  |
| 9 | Mideksa et.al (2018) | 34 | 142 | 32 |  |  | 189 | 19 | 163 | 39 |  |  |  |  |  |  |
| 10 | Shimels, T. et.al (2018) | 45 | 174 |  | 119 | 100 |  |  |  |  |  |  |  |  | 118 | 101 |
| 11 | Yigazu, D. &Desse, T. (2017) |  |  |  | 49 | 54 |  |  |  | 52 |  |  | 51 | 50 |  |  |

**Table 3:**
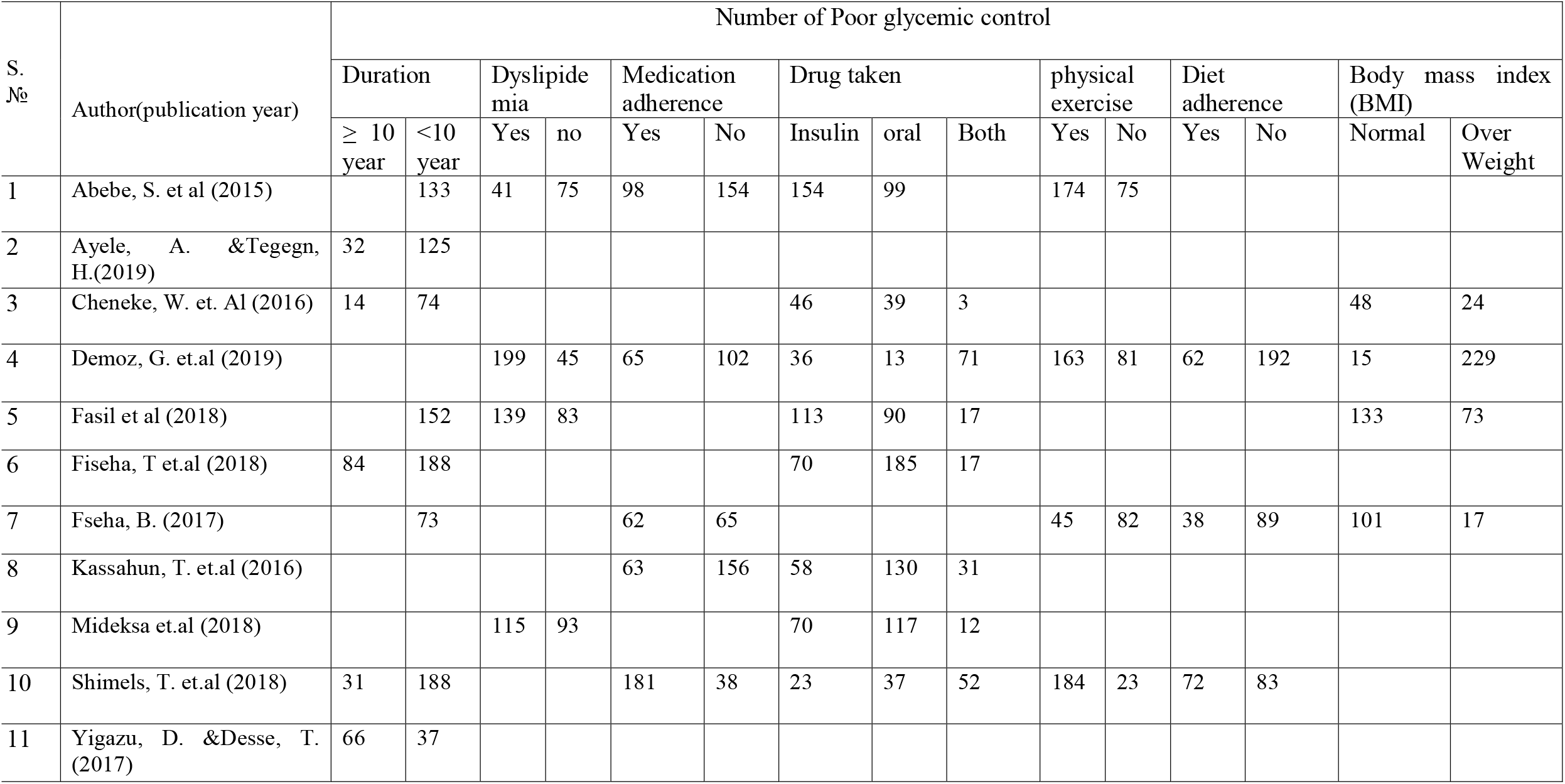
Distribution of factors associated with poor glycemic control in the selected studies in Ethiopia, 2019

### Pooled prevalence of poor glycemic control among diabetic patients

The pooled prevalence of poor glycemic control among diabetic patients in Ethiopia is 64.72% with 95% confidence interval 63.16-66.28. Heterogeneity test was checked in this analysis and it was I^2^ =73.8% indicating of no heterogeneity. Egger’s test was used to test for publication bias and t-test= −48, p-value = 0.638, hence we declare that there were no publication bias in this analysis. Figure 2

**Figure 2:**
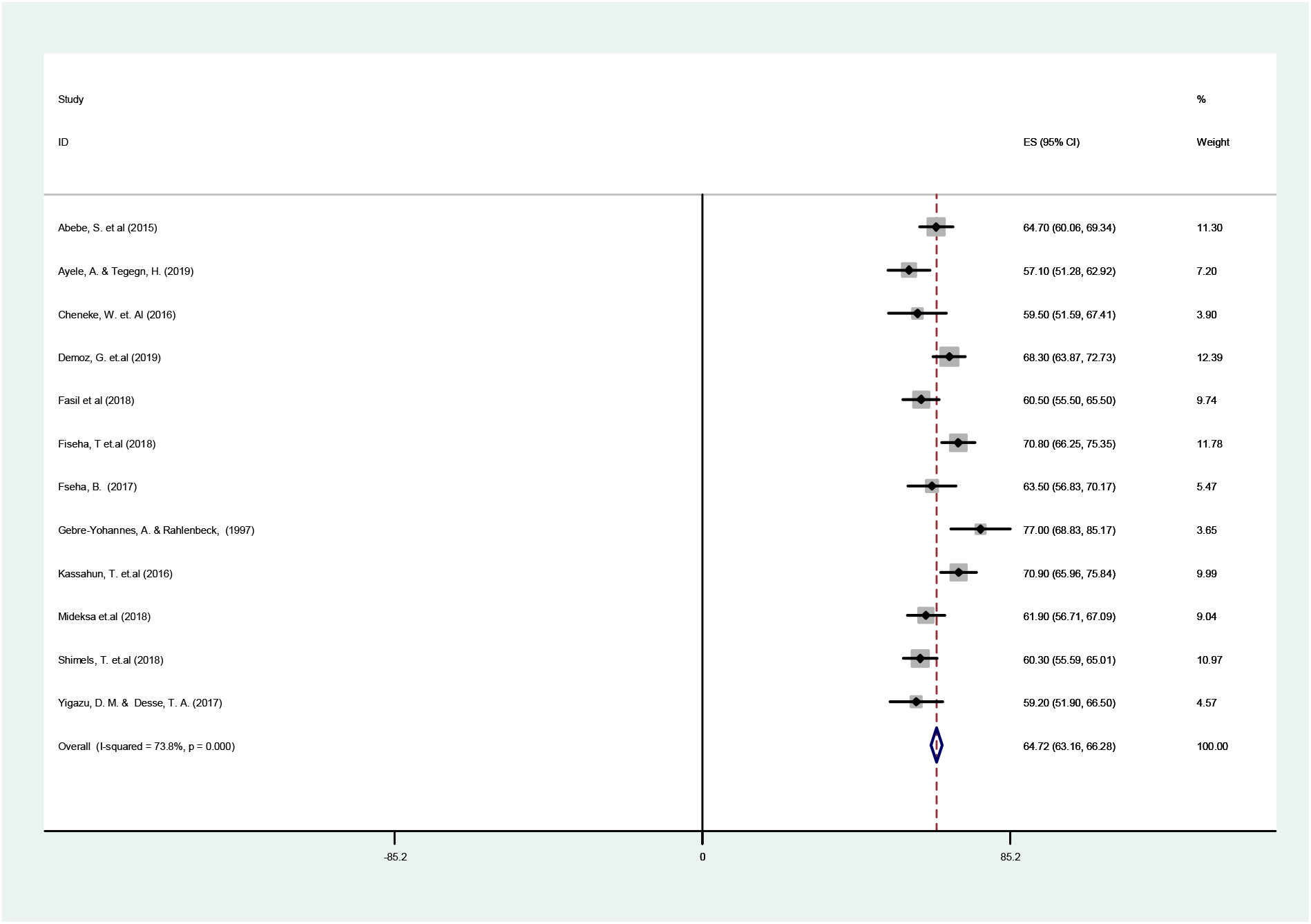
forest plot of the 12 studies assessed for the pooled prevalence of poor glycemic control among diabetic patients in Ethiopia, 2019.

#### Sensitivity test

Sensitivity test has been conducted in the quantitative meta-analysis of poor glycemic control among diabetic patients in Ethiopia for the selected studies and all of the articles were with in the 95% confidence interval. Figure 3.

**Figure 3:**
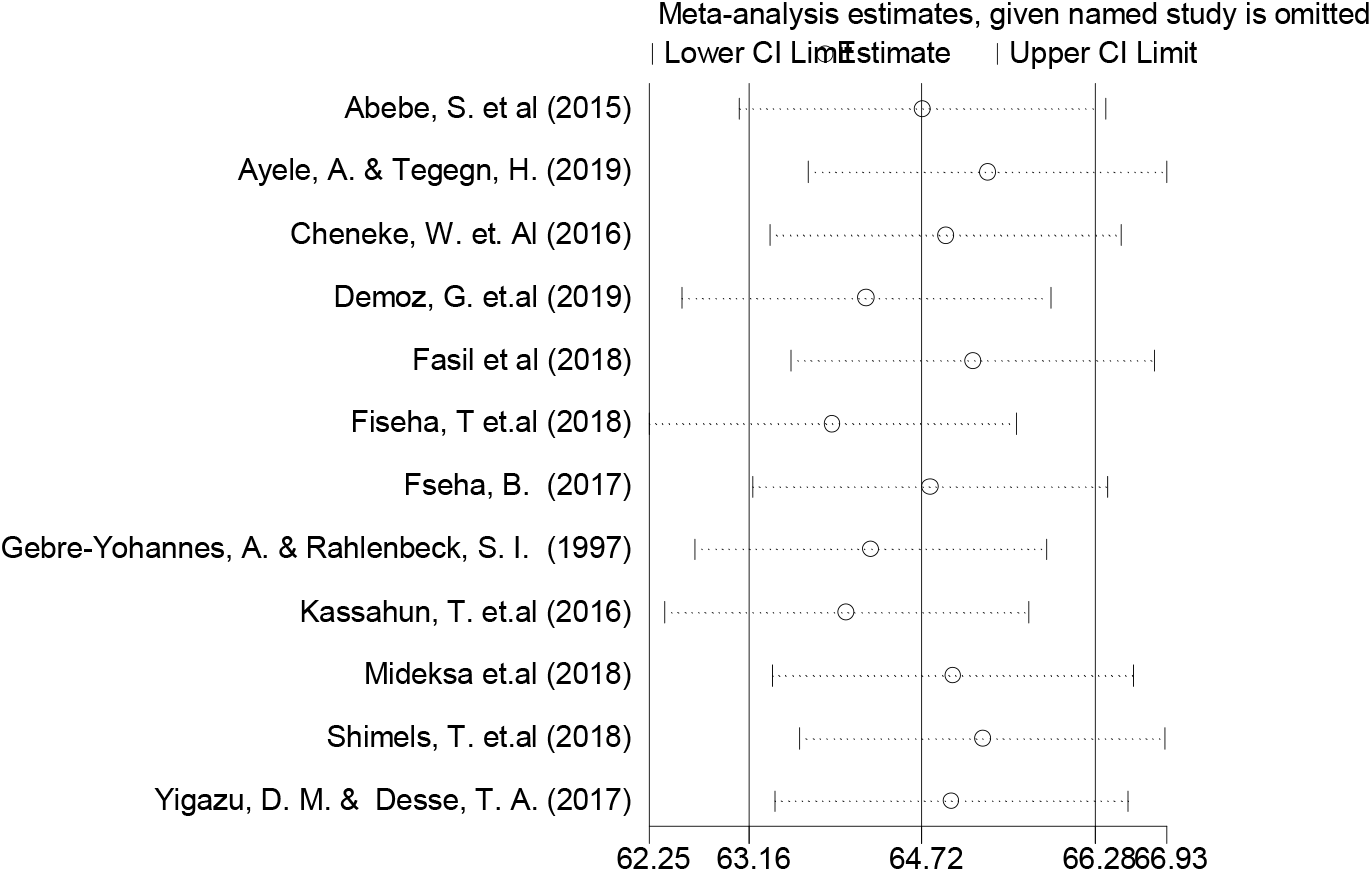
meta-analysis estimates of sensitivity test for the selected studies on poor glycemic control among diabetic patients in Ethiopia, 2019.

#### Subgroup analysis of poor glycemic control by region of the country

The sub group analysis of poor glycemic control among diabetic patients in different region of the country shows consistent and high prevalence of poor glycemic control ranging from 62.5% in Tigray region to 65.6% in Oromia region of the country. In Amhara region five studies have been included in the meta-analysis and the overall prevalence of poor glycemic control among diabetic patients was 65.18%. (Figure 4).

**Figure 4:**
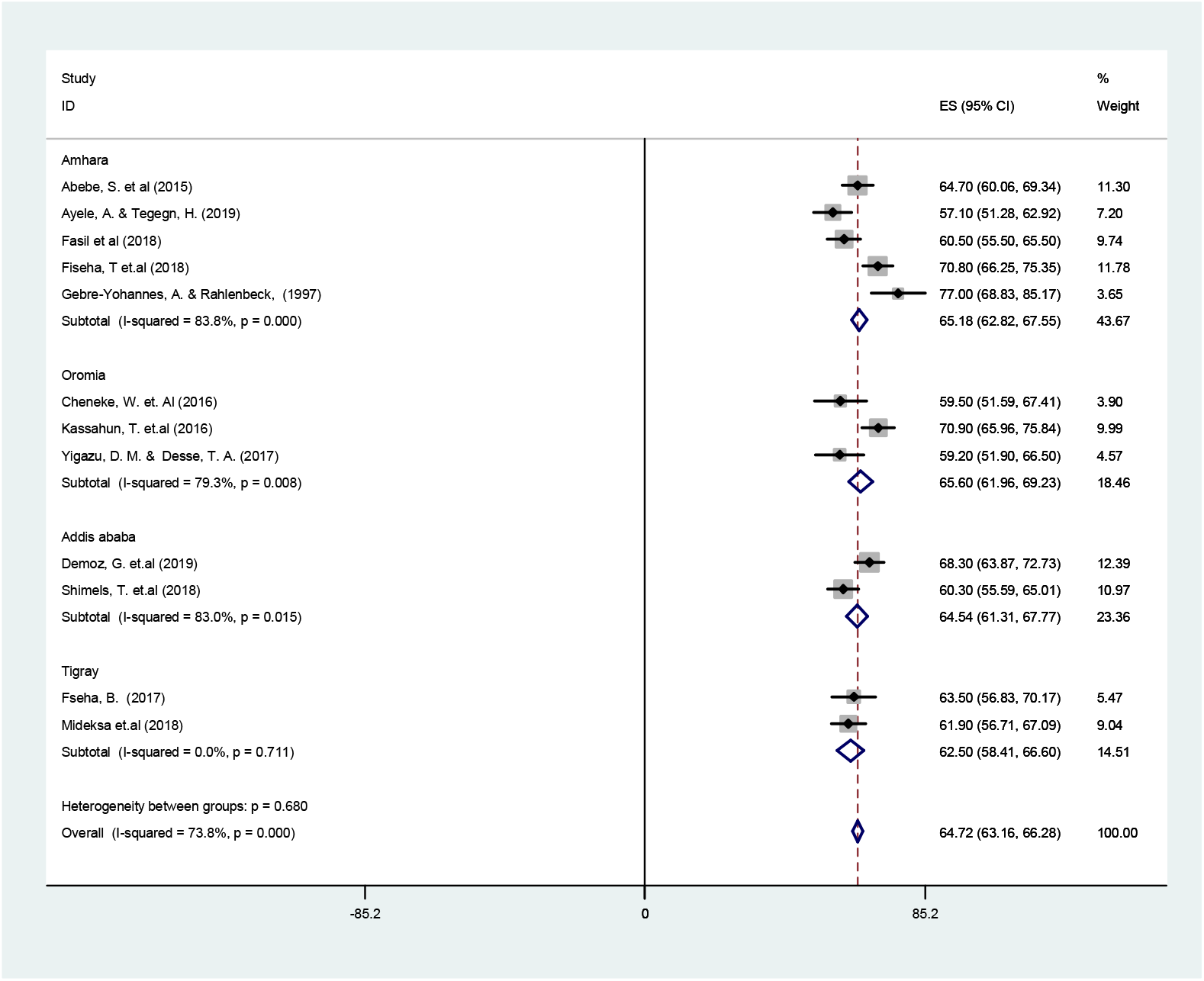
forest plot showing sub group anlysis byregion of the country of the poor glycemicin Ethiopia, 2019

#### Subgroup analysis of poor glycemic control by method of measuring blood glucose level

The subgroup analysis of poor glycemic control among diabetic patients by the fasting blood glucose (FBG) and by glycated hemoglobin (HbA1c) shows that there was no difference in the result of poor glycemic control. Eight studies has been included in quantitative meta analysis of the systemic review measured by fasting blood glucose and the overall poor glycemic control measured by FBG was 64.74%. Studies measured the poor glycemic control in the review were four and the overall poor glycemic contral was 64.68%. Figure 5

**Figure 5:**
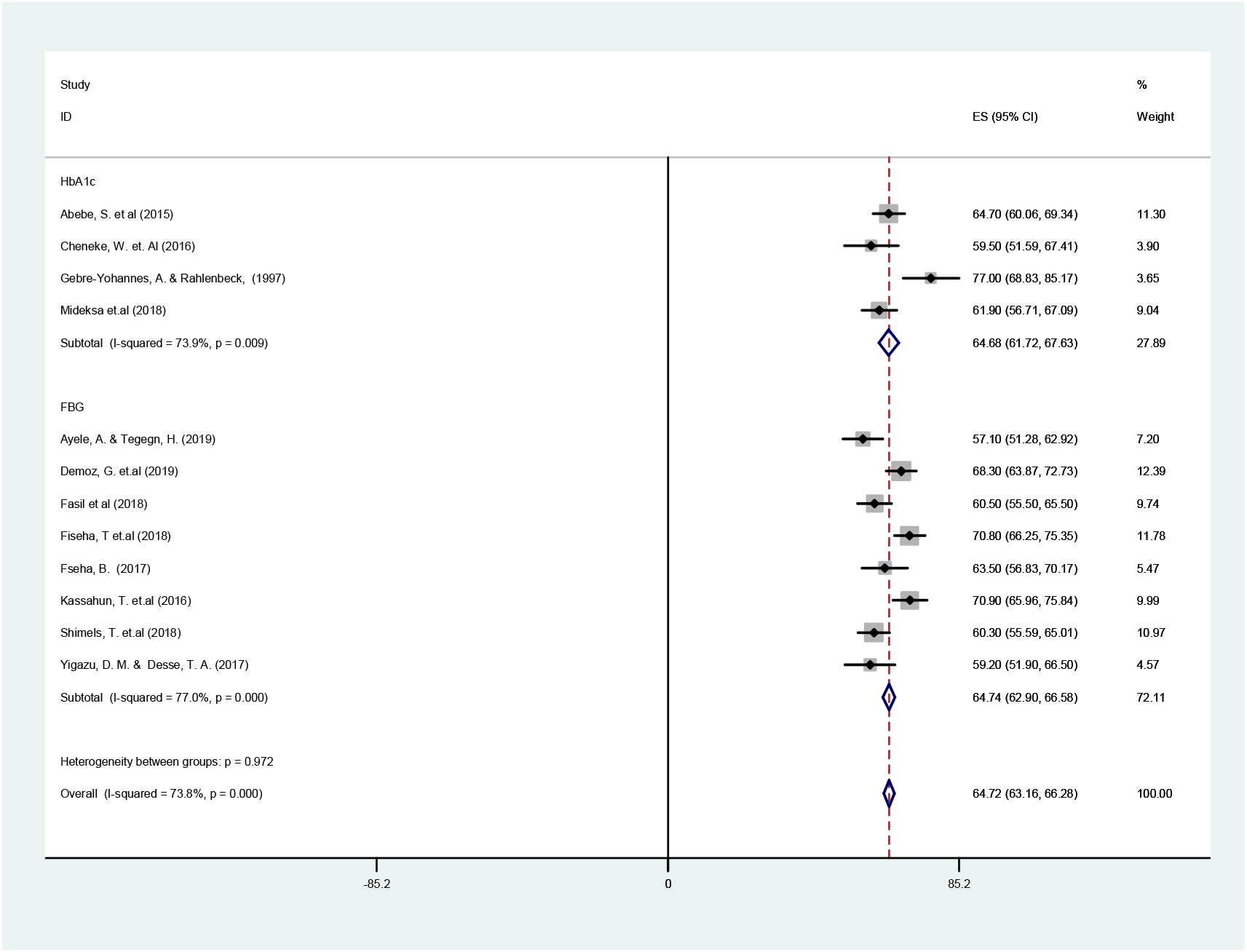
forest plot showing sub group anlysis by method of measuring blood glucose level of the poor glycemicin Ethiopia, 2019

#### Factors associated with poor glycemic control among diabetic patients

To identify factors that have significant association with poor glycemic control among diabetic patients we had conducted the quantitative meta-analysis of with random and fixed effect model based on the heterogeneity of the studies and random effect model is the more conservative method. There were a total of 14 factors investigated for poor glycemic control, among those only residence, dyslipidemia and diet adherence were significantly associated with poor glycemic control among diabetic patients in Ethiopia after conducting meta-analysis.

#### Residence

In the quantitative meta-analysis total of seven studies have been including to identify residence (rural/urban) as a significant factor associated with poor glycemic control. Fixed effect model was used because there was no heterogeneity (I^2^ =0.0%, p=0.43). Being rural residence was 1.74 times more likely to have poor glycemic control among diabetic patients as compared to urban residences. Figure 6

**Figure 6:**
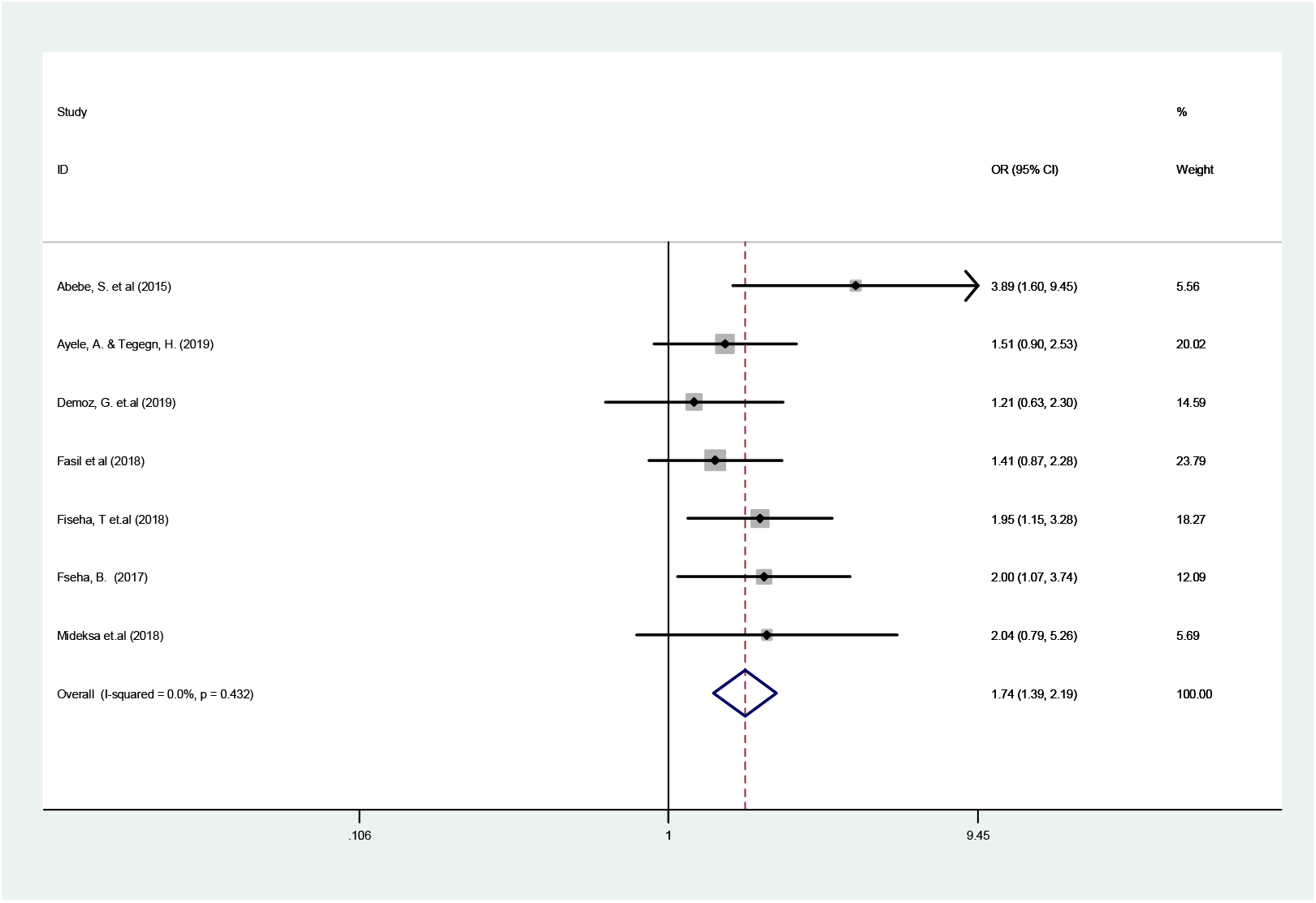
Forest plot showing the pooled significance of residence (rural/urban) with poor glycemic control among diabetic patients in Ethiopia, 2019.

#### Dyslipidemia

In the quantitative meta-analysis total of four studies have been including to identify dyslipidemia (normal lipid level/abnormal lipid level) as a significant factor associated with poor glycemic control. Random effect model was used because there was heterogeneity (I^2^ =84.3%, p=<0.001). Being dyslipidemia was 4.69 times more likely to have poor glycemic control among diabetic patients as compared to patients that had normal level of blood lipids. Figure 7

**Figure 7:**
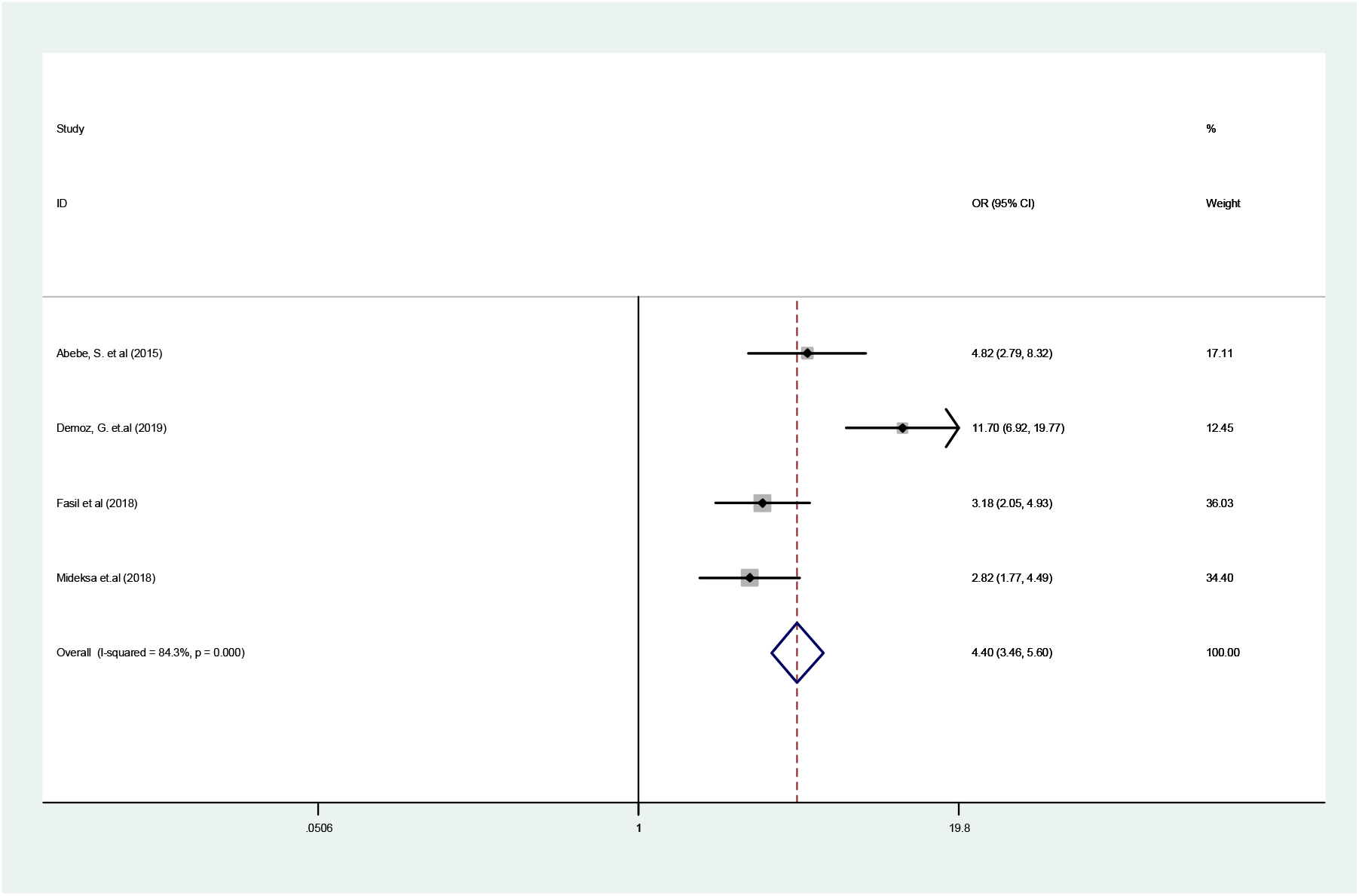
Forest plot showing the pooled significance of dyslipidemia (abnormal/normal) with poor glycemic control among diabetic patients in Ethiopia, 2019

#### Diet adherence

In the quantitative meta-analysis total of three studies have been including to identify dietary plan adherence as a significant factor associated with poor glycemic control. Random effect model was used because there was heterogeneity (I^2^ =90%, p=<0.001). Being not adhered to the dietary plan was 5.45 times more likely to have poor glycemic control among diabetic patients as compared to patients that adhere to the dietary plan. Figure 8

**Figure 8:**
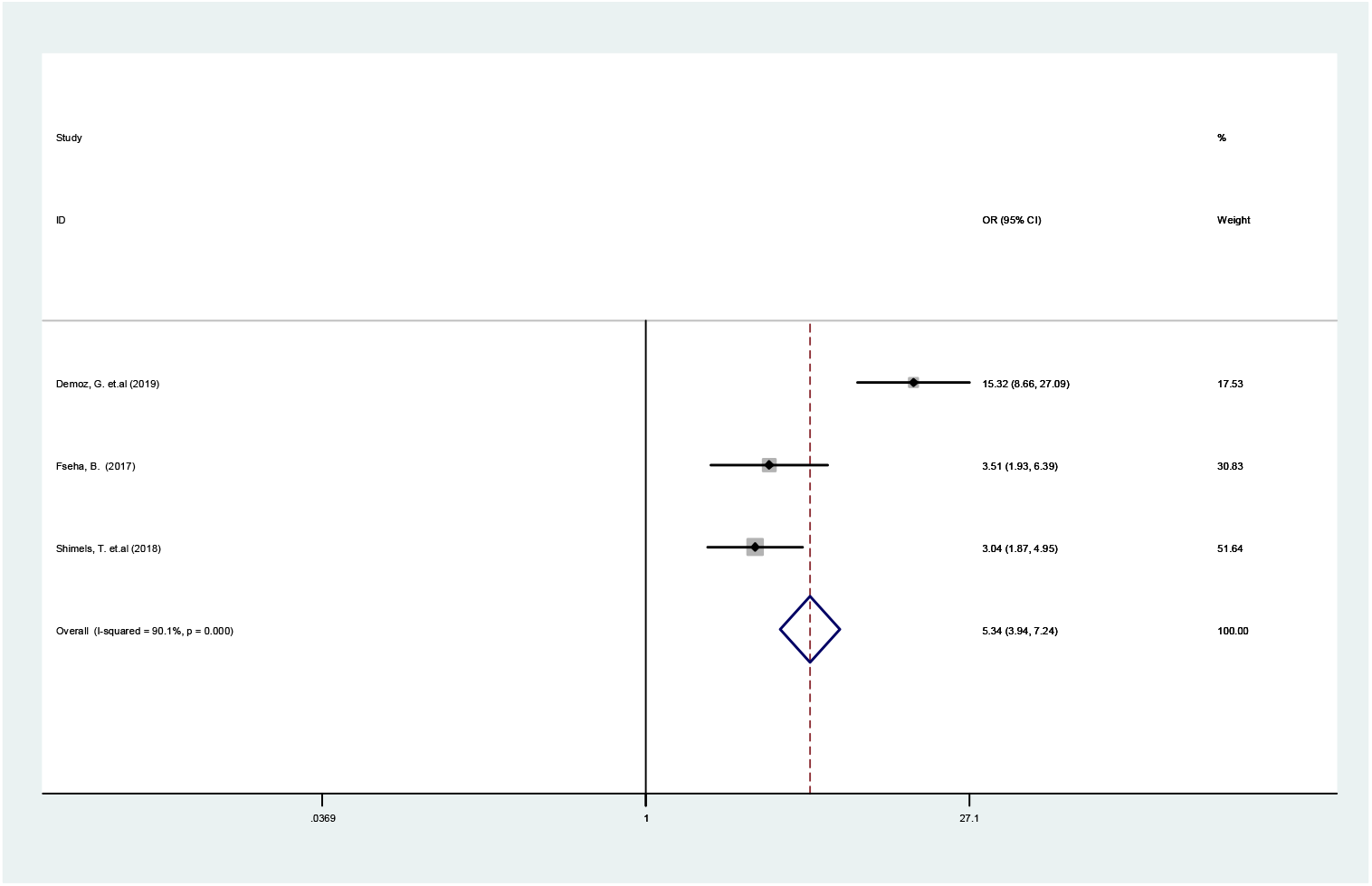
Forest plot showing the pooled significance of dietary plan adherence with poor glycemic control among diabetic patients in Ethiopia, 2019.

## Discussion

Poor glycemic control among diabetic patients is the major cause of macro vascular and micro vascular complication and lead to hospital admissions.

In the current systemic review and meta-analysis a total of twelve studies have been used to summarize the pooled prevalence of poor glycemic control and their associated factors among diabetic patients in Ethiopia. The overall pooled prevalence of poor glycemic among diabetic patients was 64.72% and being rural residence, dyslipidemia, and not adhering to dietary plan were factors that had significant association with poor glycemic control.

The result of poor glycemic control in the current study 64.72% with 95% CI (63.16 – 66.28) is consistent with study conducted in Jordan 65.1%, (9), India 63% (10),. However the current finding is higher than a study conducted in china 50.3%(12), Zimbabwe 58.2% (13), Israel 58.4%(14). This discrepancy might be due to the study population characteristics, study design and the kind of medication they were given. Besides, the pooled prevalence of poor glycemic control among diabetic patients is lower than a study conducted in Bangladish 82% (19), manipal/India 78.2%(20), 78.6%, Kenya 91.8%(21), Saudi Arabia 74.9%(22). This variation could be due to the measurement tool used for the outcome determination.

In the Meta analysis to identify significant factors associated with poor glycemic control among diabetic patient’s twelve studies has been included. Being rural residence was 1.74 times more likely to have poor glycemic control as compared to urban residence(OR=1.72, 95%CI 1.39,2.19), this result is in line with a study conducted in Bangladesh(19),. This could be explained by peoples living in rural residence might have lower awareness, and also may not have sufficient access to health facility for professional support to maintain their glycemic control as compared to urban residences(25).

In addition being dyslipidemia was 4.69 times more likely to have poor glycemic control as compared to the patients having normal lipid level (OR=4.69, 95%CI 2.51,8.71) this result is in line with a study conducted by Kakade A. et al 2018 in Mumbai/India(21), systemic review by mohammad J. et al 2018 in Arabian gulf countries(26),. This could be due to the fact that continuous entry of fatty acids in to the β-cells of the pancreas creates β-cell lipotoxicity and this causes pancreases β-cell failure follow on poor glycemic control(27–29).

Similarly finding of this study revealed being no adherent to the dietary plan was 5.45 more likely to have poor glycemic control as compared to patients that adhere to their dietary paln (OR=5.45, 95%CI 1.99,14.92) this result is in line with a study conducted by Kakade A. et al 2018 in Mumbai/India(21), systemic review by Mohammad J. et al 2018 in Arabian gulf countries(26), and Khattab M. et al. 2010 in Jordan(9). This could be due to the fact that diet is the most important treatment that can modify may aspects of the body physiology including the pathophysiology of pancreas that secretes the hormone insulin(5).

## Conclusion

The prevalence of poor glycemic control among diabetic patients was high in Ethiopia and consistent across different regions of the country. The most important factors associated with poor glycemic factor among diabetic patients were being rural residence, having dyslipidemia and not adhering to dietary plan.

## Data Availability

all the data set used in the analysis availabele with corresponding author and can deliver on request.

## Abbreviations

FBG: Fasting blood glucose
HbA1c: Glycated hemoglobin
BMI: Body Mass Index.

## Declarations

### Ethics approval and consent to participate

N/A

### Consent for publication

Not applicable

### Availability of data and material

The datasets used and/or analyzed during the current study are available from the corresponding author on reasonable request.

### Authors contributions

BFT participated in the preparation of the design of the study, data analysis, write up and preparing daft of the manuscript and critically reviewing. Similarly AKB and GGW, participated in the design of the study, data analysis and presentation, reviewing of the manuscript. All authors read, accepted and approved the final manuscript

